# Patient Perceptions of Student Involvement and Satisfaction in a Student-Run Free Clinic Serving People Experiencing Homelessness

**DOI:** 10.64898/2026.10.13.26365447

**Authors:** Mackenzie K. Joe, Spoorthi Kamepalli, Kevin Hoffman, Alexa Cumming, Lance Babcock, Dillon Noltensmeyer, Shragvi Balaji, Cathryn Bigham, Benjamin T. King, Michael Altman, Malford T. Pillow, Victoria McCurry, Dana L. Clark

**Author notes:** **Funding:** None. **Patient and public involvement:** Patients were not involved in the design, conduct, reporting, or dissemination planning of this study. Patients participated by voluntarily completing anonymous post-visit satisfaction surveys. The survey instrument is provided in Supplemental Table S1. This study is reported in accordance with the Strengthening the Reporting of Observational Studies in Epidemiology (STROBE) guidelines for cross-sectional studies.

## Abstract

**Background:** Patient satisfaction is a key measure of healthcare quality among people experiencing homelessness. Student-run free clinics (SRFCs) provide care to underserved populations, but the role of student involvement in patient satisfaction remains unclear.

**Methods:** We conducted a cross-sectional study of patients receiving care at an SRFC. From February 2024 to February 2025, patients completed post-visit surveys assessing satisfaction that were evaluated using regression analyses.

**Results:** Eighty-eight patients completed surveys, reporting high overall satisfaction (mean 9.26/10). In separate linear regression models adjusted for age, gender, and race, greater perceived benefit of student involvement was associated with higher satisfaction (β = .298, p =.037), whereas perceiving the visit as too long was associated with lower satisfaction (β = −.285, p = .019).

**Conclusion:** Greater perceived benefit of student involvement was associated with higher patient satisfaction, whereas longer perceived visit duration was associated with lower satisfaction. These findings highlight the importance of balancing educational opportunities with efficient workflows in SRFCs.

## Manuscript

Patient satisfaction is an important indicator of healthcare quality and is associated with treatment adherence, continuity of care, and patient engagement.^1,2,3^ Although patient satisfaction is influenced by multiple factors, including expectations, communication, and perceived quality of care, it remains a valuable patient-reported outcome for evaluating healthcare delivery and guiding quality improvement initiatives.^3,4,5^ These measures are particularly relevant in underserved populations, where trust and ongoing engagement with healthcare systems are critical.^2,6^

Student-run free clinics (SRFCs) occupy a unique position at the intersection of patient care, medical education, and community advocacy, providing healthcare to underserved populations while serving as important educational settings for future physicians.^7^ Although SRFCs consistently report high levels of patient satisfaction, the specific contribution of medical student involvement remains less understood.^8-10^ A recent meta-analysis by Brookshire et al. found that overall patient satisfaction was generally comparable between SRFCs and traditional clinical settings; however, differences emerged in specific domains of care, suggesting that medical students may influence aspects of the patient experience rather than overall satisfaction alone.^11,12^ These findings highlight the need to better understand which components of student involvement are associated with patient satisfaction, particularly among people experiencing homelessness.

For people experiencing homelessness, patient satisfaction may be particularly important because previous healthcare experiences frequently involve stigma, discrimination, and barriers to care.^13,14^ Studies have demonstrated that individuals experiencing homelessness often report feeling unwelcome, rushed, or dismissed during healthcare encounters, contributing to reduced trust, delayed healthcare seeking, and poorer continuity of care.^15,16^ Consequently, understanding which aspects of healthcare encounters contribute to positive patient experiences may help clinics better address the needs of this vulnerable population.^6,17^

Although patient satisfaction has been evaluated in several SRFCs, little is known about which aspects of care are associated with patient satisfaction among people experiencing homelessness.^12^ The Houston Outreach Medicine Education and Social Services (HOMES) clinic is Healthcare for the Homeless-Houston’s (HHH) student-run clinic and operates entirely under the umbrella of HHH. The HOMES clinic is adjoined to the Beacon, which serves individuals experiencing homelessness by providing daily meals, showers, civil legal aid, counseling, and mentoring. HOMES operates as an urgent care-style clinic where patients can return for episodic care and are referred to primary care and specialty services, such as dermatology or ophthalmology, for ongoing, longitudinal care. The clinic is staffed by students from several local medical schools and pharmacy schools, providing interdisciplinary care to individuals experiencing homelessness. While previous studies have primarily focused on overall patient satisfaction or educational outcomes within SRFCs, few have examined how perceived student involvement, patient-centered communication, and other visit characteristics are associated with patient satisfaction in this population.^11^ Understanding these relationships may inform quality improvement efforts and the continued development of student-run models of care.^7^

The objective of this study was to evaluate patient satisfaction among individuals receiving care through the HOMES clinic and to identify factors associated with overall satisfaction. We hypothesized that greater perceived benefit of student involvement and patient-centered communication would be associated with higher patient satisfaction. We additionally sought to identify which aspects of the patient experience were most strongly associated with overall visit satisfaction to guide future quality improvement initiatives.

## Methods

### Population Studied

This cross-sectional study evaluated patient satisfaction among individuals receiving care at the HOMES clinic. Between February 2024 and February 2025, all patients attending the clinic were offered the opportunity to complete a voluntary post-visit satisfaction survey immediately following their clinical encounter. No names or other direct identifiers were collected. The clinic typically serves approximately 2–6 patients per week. No formal sample size calculation was performed; the study included all 88 completed surveys collected during the study period.

Surveys were administered by student volunteers, who introduced the survey, confirmed that patients understood it, agreed to participate through verbal consent, read items aloud as needed, and assisted patients in completing a standardized REDCap form on IRB-approved tablet devices. The patient survey included demographic items, a 10-point overall satisfaction measure, 5-point and 7-point Likert-type items assessing aspects of the patient experience, and optional free-text responses. Survey domains included perceived benefit of student involvement, patient-centered communication (including listening, understandable care plans, helpfulness of the after-visit summaries, and discussion of barriers to care), accessibility, care planning, and barriers to care. Survey items included in the quantitative analyses, including response scales and scoring, are provided in Supplemental Table S1.

The primary outcome was overall patient satisfaction. Predictor variables included demographic characteristics, perceived benefit of student involvement, visit length, care planning, helpfulness of an after-visit summary, barriers to care, and accessibility measures.

### Statistical Analysis

Descriptive statistics summarize participant characteristics and survey responses. Numeric ratings were summarized using means and standard deviations; categorical characteristics were summarized using counts and percentages. Likert-type items were treated as continuous variables. The Kruskal–Wallis H test was used to compare overall satisfaction across categorical demographic variables. Bivariate linear regression analyses were performed to evaluate associations between individual survey items and overall satisfaction. Communication-related items were examined in bivariate models only.

Perceived benefit of student involvement and perceived visit length were evaluated in separate multivariable ordinary least squares (OLS) regression models, each adjusted for age, gender, and race. Overall satisfaction was analyzed as a continuous outcome to facilitate interpretation of regression coefficients, consistent with prior analyses of patient satisfaction measures. Because the overall satisfaction outcome demonstrated a marked ceiling effect, regression analyses were interpreted cautiously. Analyses were performed using an available-case approach because survey items were optional; missing responses were not imputed, and sample sizes therefore varied across analyses according to the variables required for each analysis.

All analyses were performed using Python 3.13.5 with the statsmodels and patsy libraries in Visual Studio Code. This study is reported in accordance with the Strengthening the Reporting of Observational Studies in Epidemiology (STROBE) guideline for cross-sectional studies.^24^ The study was approved and monitored by the Baylor College of Medicine Institutional Review Board (H-53843).

### Patient and Public Involvement

Patients and the public were not involved in the design, conduct, or reporting of this study.

## Results

### Survey Results

A total of 88 patients completed the patient satisfaction survey following care at the HOMES clinic. The number of patients approached was not recorded. Participant demographic characteristics are summarized in Table 1.

**Table 1.** Participant characteristics of survey respondents at the HOMES clinic February 2024-February 2025.

| Characteristic | Value |
| --- | --- |
| Total Participants | 88 |
| Age, mean (SD) | 52.4 (11.3) |
| Female, n/N (%) | 11/87 (12.6%) |
| Black, n/N (%) | 49/87 (56.3%) |
| White, n/N (%) | 28/87 (32.2%) |
| Other race, n/N (%) | 10/87 (11.5%) |

Statistics reflect available responses: age, n = 81; gender and race, n = 87 each. Other race includes American Indian, more than one race, and not listed. SD, standard deviation.

Overall patient satisfaction was high, with a mean overall satisfaction score of 9.26 (SD = 1.9) on the 10-point scale; 81.4% of participants selected the highest possible rating. Participants reported high ratings of perceived benefit of student involvement (mean = 9.2, SD = 1.6).

Respondents strongly agreed that the care team listened to their concerns, provided an easy-to-understand care plan, provided a helpful after-visit summary, and discussed barriers to care (all mean scores 6.2–6.6 on the 7-point scale) (Table 2). The mean perceived visit-length rating was 3.3 (SD 1.4) on the 5-point scale.

**Table 2.**
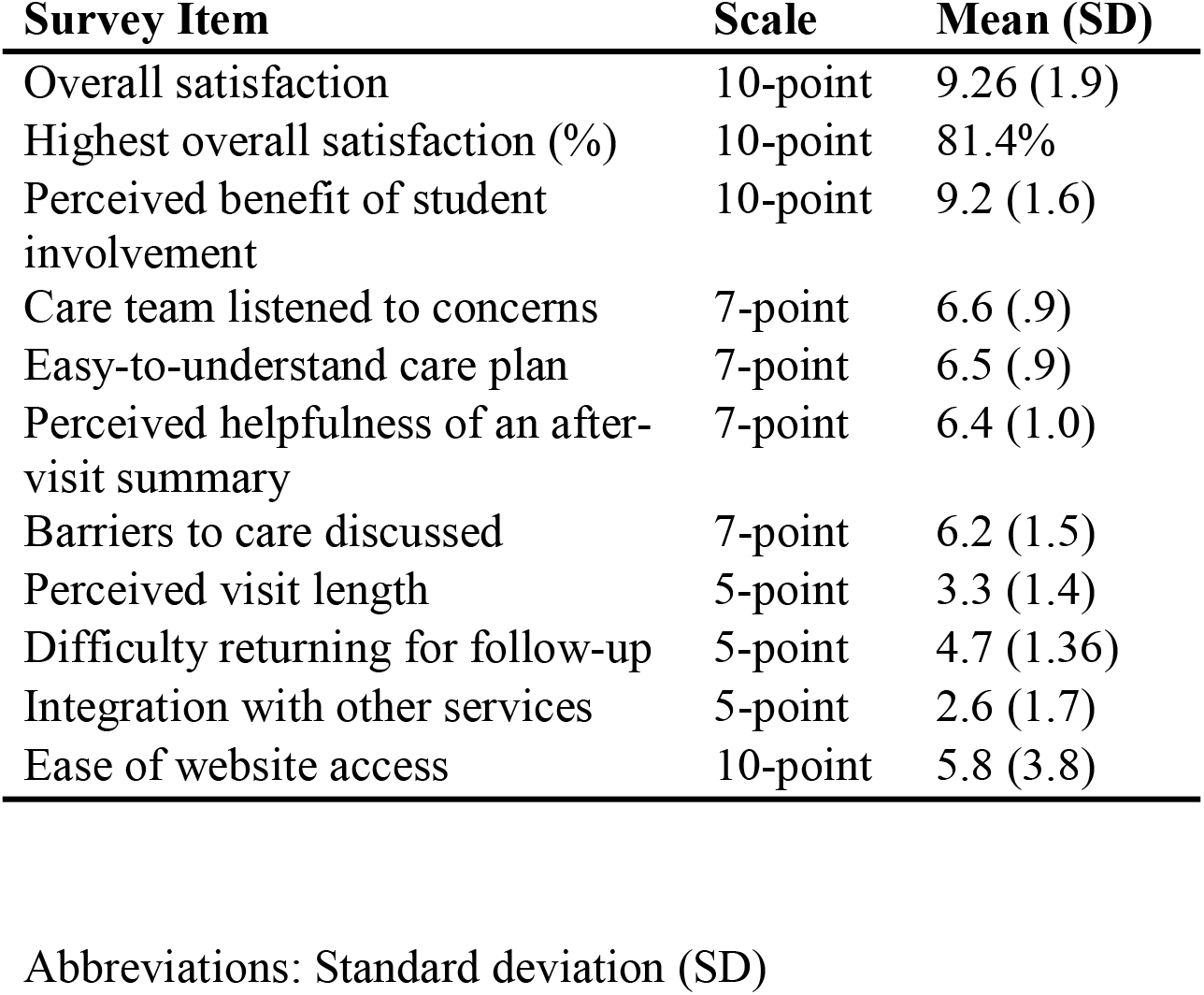
Patient satisfaction survey responses by survey item, scale, mean, and standard.

| **Survey Item** | **Scale** | **Mean (SD)** |
| --- | --- | --- |
| Overall satisfaction | 10-point | 9.26 (1.9) |
| Highest overall satisfaction (%) | 10-point | 81.4% |
| Perceived benefit of student involvement | 10-point | 9.2 (1.6) |
| Care team listened to concerns | 7-point | 6.6 (.9) |
| Easy-to-understand care plan | 7-point | 6.5 (.9) |
| Perceived helpfulness of an after-visit summary | 7-point | 6.4 (1.0) |
| Barriers to care discussed | 7-point | 6.2 (1.5) |
| Perceived visit length | 5-point | 3.3 (1.4) |
| Difficulty returning for follow-up | 5-point | 4.7 (1.36) |
| Integration with other services | 5-point | 2.6 (1.7) |
| Ease of website access | 10-point | 5.8 (3.8) |

Abbreviations: Standard deviation (SD)

Patients reported greater difficulty returning for follow-up visits (mean = 4.7, SD = 1.36), whereas integrating the clinic visit with other available services was rated relatively easy (mean = 2.6, SD = 1.7). Participants also reported moderate ease of accessing online resources (mean = 5.8, SD = 3.8) (Table 2).

### Satisfaction Associations

Bivariate linear regression identified greater perceived benefit of student involvement (β = .571, P < .001, n = 85) and care team listening (β = .752, P < .001, n = 86) as significantly associated with higher overall satisfaction. Positive associations were also observed for an easy-to-understand care plan (β = .683, P = .003, n = 85), perceived helpfulness of the after-visit summary (β = .698, P < .001, n = 83), and discussion of barriers to care (β = .411, P = .001, n = 83). Because the majority of participants selected the highest possible satisfaction rating, these findings should be interpreted in the context of a pronounced ceiling effect.

In separate multivariable linear regression models adjusted for age, gender, and race, greater perceived benefit of student involvement remained significantly associated with higher overall satisfaction (β = .298, P = .037, n = 78), whereas perceiving the visit as too long was associated with lower satisfaction (β = −.285, P = .019, n = 78) (Table 3). Demographic characteristics were not significantly associated with patient satisfaction (Figure 1).

**Table 3.** Adjusted linear regression results for overall patient satisfaction controlling for age, gender, and race in ordinary least squares regression.

| **Predictor** | **$\beta$** | **p-value** | **n** |
| --- | --- | --- | --- |
| Perceived benefit of student involvement | .298 | .037 | 78 |
| Visit perceived as too long | -.285 | .019 | 78 |

**Figure 1:**
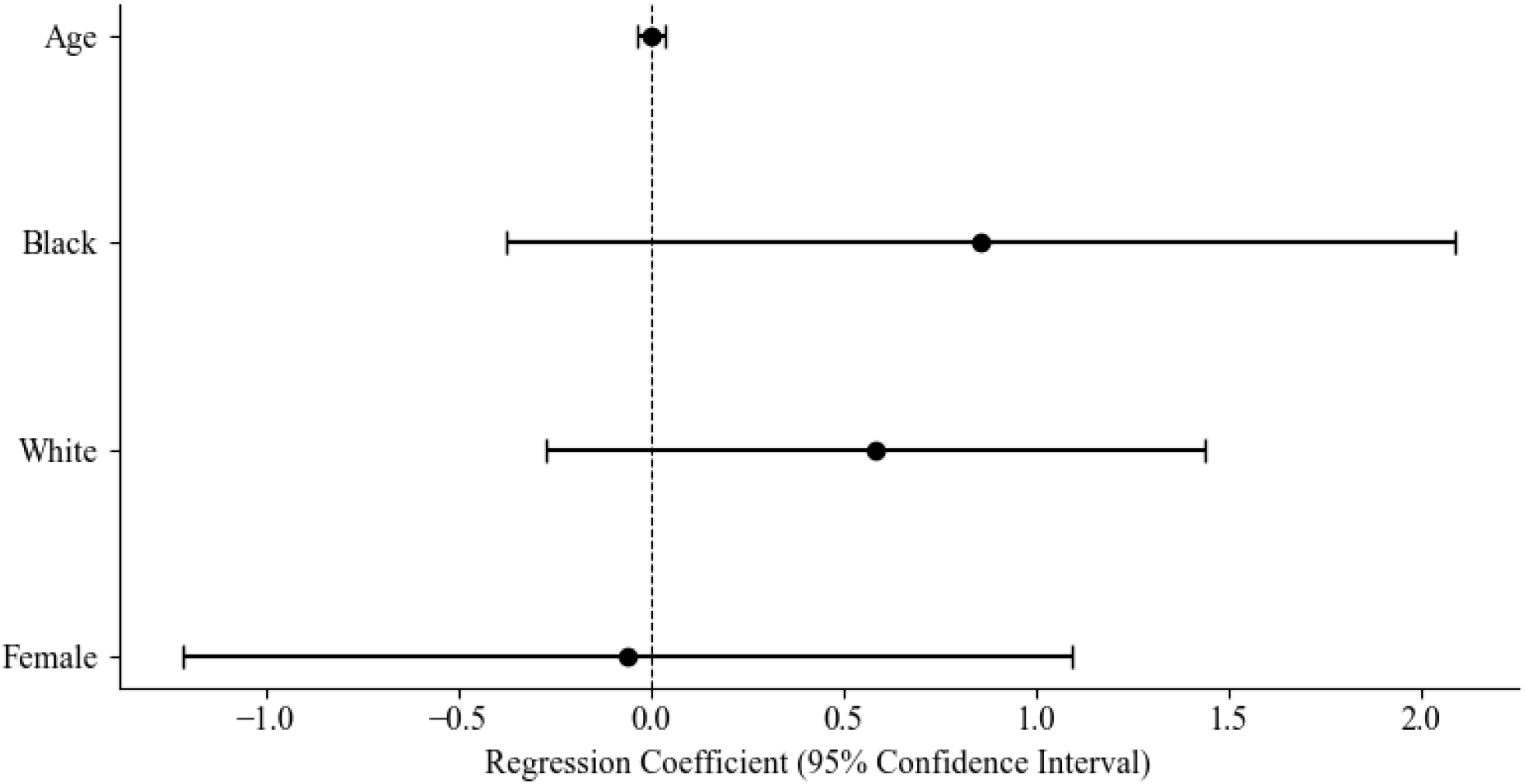
Adjusted associations between participant characteristics and overall patient satisfaction

## Discussion

This study evaluated patient satisfaction at the HOMES student-run clinic serving people experiencing homelessness. Overall patient satisfaction was high across multiple domains, and greater perceived benefit of student involvement was associated with higher patient satisfaction. Patient-centered communication—including listening to patient concerns, providing understandable care plans, and discussing barriers to care—was also associated with higher satisfaction, whereas longer perceived visit duration was associated with lower satisfaction. Together, these findings support continued evaluation of patient-centered communication strategies within SRFCs serving vulnerable populations.

Student involvement was associated with higher patient satisfaction. Patients may perceive greater trainee involvement positively rather than detracting from their care experience. ^8, 9, 18^ This extends the growing literature on SRFCs by evaluating patients’ perceived benefits of student involvement within a clinic dedicated to serving people experiencing homelessness. In addition to providing meaningful educational experiences for trainees, SRFCs may foster patient-centered communication among underserved populations.^7, 19, 20^

Communication-related factors—including listening to patient concerns, providing understandable care plans, after-visit summaries, and discussing barriers to care—were associated with higher patient satisfaction, underscoring the importance of interpersonal communication in building trust among people experiencing homelessness. These findings are consistent with prior literature demonstrating that communication, empathy, and individualized care are among the strongest determinants of patient satisfaction.^21, 22^ Although this study cannot determine which aspects of these interactions were most influential, the results reinforce the importance of patient-centered communication in caring for underserved populations. Communication-related associations were examined in bivariate models only and were not adjusted for age, gender, or race.

Longer perceived visit duration was associated with lower patient satisfaction despite the positive association between student involvement and satisfaction. This finding highlights an important challenge for SRFCs. For many people experiencing homelessness, SRFCs may be among the few sources of physician care, helping bridge a substantial gap in access to the healthcare system.^7^ SRFCs are also educational settings, and the added time students spend on histories, examinations, and case presentations may lengthen visits. Patients managing competing daily needs while experiencing homelessness may be especially sensitive to long waits.^6,15^ At the same time, longer visits may allow for more comprehensive counseling and a fuller discussion of barriers to care. Future studies should evaluate strategies that preserve the interpersonal benefits of student involvement while improving clinic efficiency.^11,23^

This study has several methodological strengths. Surveys were administered immediately after clinic visits, reducing recall bias and capturing patients’ experiences while the encounter was still recent. Survey data were collected over multiple months and clinic sessions, and the HOMES Clinic is staffed by students from three medical schools, increasing the diversity of trainee experiences represented.

Several limitations warrant consideration. First, the cross-sectional design precludes causal inference. Second, while all patients were offered the opportunity to complete the survey, the total number of eligible patients was not systematically recorded, which prevented calculation of a response rate, limited assessment of nonresponse bias, and, because surveys did not collect identifiers, precluded identification of repeat respondents. Additionally, although immediate administration reduced recall bias, self-reported responses collected right after the visit, including perceived benefit of student involvement, may have been influenced by participants’ overall satisfaction with their visit. Because surveys were administered by student volunteers affiliated with the clinic, social desirability bias may also have contributed to the high satisfaction ratings. The single-center design and modest sample size may limit generalizability. Overall satisfaction demonstrated a pronounced ceiling effect, limiting response variability, and warranting cautious interpretation of the regression analyses. Furthermore, multiple hypothesis tests were performed without adjustment for multiple comparisons; therefore, statistically significant findings should be interpreted as exploratory. Finally, patient experiences may also have varied across individual student providers, and the absence of a comparison group precluded direct comparison with traditional healthcare settings.

Individuals experiencing homelessness receiving care through the HOMES student-run clinic reported high levels of satisfaction, with perceived student involvement and patient-centered communication emerging as factors associated with positive patient experiences. These findings suggest that SRFCs such as the HOMES clinic can deliver positive patient experiences while serving as valuable educational environments for medical trainees.

## Conclusion

Individuals experiencing homelessness receiving care reported high levels of satisfaction. Greater perceived benefit of student involvement and patient-centered communication were associated with higher overall satisfaction, whereas longer perceived visit duration was associated with lower satisfaction. These findings support the continued role of student-run free clinics as educational environments capable of delivering positive patient experiences while highlighting opportunities to optimize clinic efficiency. Continued evaluation of patient experience, clinical outcomes, and trainee development will help improve care delivery and inform best practices for SRFCs serving underserved populations. Future studies should evaluate whether positive patient experiences at student-run free clinics translate into improved longitudinal engagement with healthcare and community support services.

## Data Availability Statement

The de-identified data underlying this study are available from the corresponding author upon reasonable request, subject to approval by the Baylor College of Medicine Institutional Review Board (H-53843). The survey instrument is provided in Supplemental Table S1.

## Acknowledgements

The authors would like to express their sincere gratitude to Healthcare for the Homeless–Houston and The Beacon for their continued partnership and support of the HOMES Clinic. We are especially grateful to the patients who participated in this study and shared their experiences. We also thank Carly Brown and Kathryn Rogers of Healthcare for the Homeless–Houston for their invaluable assistance and contributions to this project.

## References

1. Cleary PD, McNeil BJ. Patient satisfaction as an indicator of quality care. Inquiry. 1988;25:25–36.

2. Dubina MI, O’Neill JL, Feldman SR. Effect of patient satisfaction on outcomes of care. Expert Rev Pharmacoecon Outcomes Res. 2009;9(5):393–395.

3. Ferreira DC, Vieira I, Pedro MI, Caldas P, Varela M. Patient satisfaction with healthcare services and the techniques used for its assessment: a systematic literature review and bibliometric analysis. Healthcare (Basel). 2023;11(5):639. doi:10.3390/healthcare11050639

4. Sitzia J, Wood N. Patient satisfaction: a review of issues and concepts. Soc Sci Med. 1997;45(12):1829–1843.

5. Hudak PL, Wright JG. The characteristics of patient satisfaction measures. Spine (Phila Pa 1976). 2000;25(24):3167–3177.

6. Wen CK, Hudak PL, Hwang SW. Homeless people’s perceptions of welcomeness and unwelcomeness in healthcare encounters. J Gen Intern Med. 2007;22(7):1011–1017.

7. Rupert DD, Alvarez GV, Burdge EJ, Nahvi RJ, Schell SM, Faustino FL. Student-run free clinics stand at a critical junction between undergraduate medical education, clinical care, and advocacy. Acad Med. 2022;97(6):824–831. doi:10.1097/ACM.0000000000004542

8. Lu KB, Thiel B, Atkins CA, Desai A, Botwin A, Povlow MR, Simms-Cendan J, Pasarica M. Satisfaction with healthcare received at an interprofessional student-run free clinic: invested in training the next generation of healthcare professionals. Cureus. 2018;10(3):e2282. doi:10.7759/cureus.2282

9. Ellett JD, Campbell JA, Gonsalves WC. Patient satisfaction in a student-run free medical clinic. Fam Med. 2010;42(1):16–18.

10. Clark JS, Bollaert A, Sills SO, Clark JH, Norris D. Patient perception of care received by students at the Jackson Free Clinic. J Miss State Med Assoc. 2014;55(4):113–118.

11. Brookshire N, Byrne JM, Mlinar T, et al. The impact of student involvement on patient satisfaction in free clinics: a meta-analysis. J Public Health. 2026.

12. Asanad K, Zheng J, Chan-Golston A, Tam E, Bhetraratana M, Lan CW, Zhao M, Abdi R, Abdi F, Vasti E, Prelip ML. Assessing quality of care through client satisfaction at an interprofessional student-run free clinic. J Interprof Care. 2018;32(2):203–210. doi:10.1080/13561820.2017.1395827

13. Reilly J, Ho I, Williamson A. A systematic review of the effect of stigma on the health of people experiencing homelessness. Health Soc Care Community. 2022;30(6):2128–2141.

14. Canham SL, et al. A scoping review of the experiences and outcomes of stigma and discrimination towards persons experiencing homelessness. Health Soc Care Community. 2024;2024:2060619.

15. Rae BE, Rees S. The perceptions of homeless people regarding their healthcare needs and experiences of receiving health care. J Adv Nurs. 2015;71(9):2096–2107.

16. O’Toole TP, Johnson EE, Borgia M, Rose J, Tailor A, Kane V. Needing primary care but not getting it: the role of trust, stigma and organizational obstacles reported by homeless veterans. J Health Care Poor Underserved. 2015;26(3):1019–1031.

17. McCabe S, Macnee CL, Anderson MK. Homeless patients’ experience of satisfaction with care. Arch Psychiatr Nurs. 2001;15(2):78–85.

18. Fröberg M, Leanderson C, Fläckman B, Hedman-Lagerlöf E, Björklund K, Nilsson GH, Stenfors T. Experiences of a student-run clinic in primary care: a mixed-method study with students, patients and supervisors. Scand J Prim Health Care. 2018;36(1):36–46. doi:10.1080/02813432.2018.1426143

19. Clark DL, Melillo A, Wallace D, Pierrel S, Buck DS. A multidisciplinary, learner-centered, student-run clinic for the homeless. Fam Med. 2003;35(6):394–397.

20. Meah YS, Smith EL, Thomas DC. Student-run health clinic: novel arena to educate medical students on systems-based practice. Mt Sinai J Med. 2009;76(4):344–356. doi:10.1002/msj.20128

21. Wolf DM, Lehman L, Quinlin R, et al. Effect of patient-centered care on patient satisfaction and quality of care. J Nurs Care Qual. 2008;23(4):316–321.

22. Batbaatar E, Dorjdagva J, Luvsannyam A, Savino MM, Amenta P. Determinants of patient satisfaction: a systematic review. Perspect Public Health. 2017;137(2):89–101.

23. Cabrera-Barona P, Blaschke T, Kienberger S. Explaining accessibility and satisfaction related to healthcare: a mixed-methods approach. Soc Indic Res. 2017;133(2):719–739.

24. von Elm E, Altman DG, Egger M, Pocock SJ, G√∏tzsche PC, Vandenbroucke JP. The Strengthening the Reporting of Observational Studies in Epidemiology (STROBE) statement: guidelines for reporting observational studies. Lancet. 2007;370(9596):1453–1457.

