## Supplemental Table 1 for "Patient Perceptions of Student Involvement and Satisfaction in a Student-Run Free Clinic Serving People Experiencing Homelessness"

**Supplementary Table S1. Patient Satisfaction Survey Items Included in Quantitative Analysis**

| **Survey Item** | **Response Scale** | **Response Anchors** |
| --- | --- | --- |
| **Participant Characteristics** | | |
| Age | Continuous (years) | |
| Gender | Male, Female, Not Listed, Prefer Not to Say | |
| Race/Ethnicity | Black, White, American Indian, Asian/Pacific Islander, More than one race, Not Listed | |
| **Overall Patient Experience** | | |
| Overall, how satisfied were you with your visit at HOMES Clinic? | 10-point Likert scale | 1 = Lowest satisfaction  10 = Highest satisfaction |
| **Student Involvement** | | |
| On a scale of 1–10, how did student involvement in your clinic visit impact your care? | 10-point Likert scale | 1 = Significantly worsened my care  10 = Significantly improved my care |
| **Communication** | | |
| My care team listened to my concerns and addressed them during my visit. | 7-point Likert scale | 1 = Strongly disagree  7 = Strongly agree |
| The care plan from my team was easy to understand and follow. | 7-point Likert scale | 1 = Strongly disagree  7 = Strongly agree |
| The After Visit Summary was helpful in reminding me what I needed to do after my appointment. | 7-point Likert scale | 1 = Strongly disagree  7 = Strongly agree |
| **Patient-Centered Care** | | |
| My healthcare team discussed or considered my barriers to care when developing my care plan. | 7-point Likert scale | 1 = Strongly disagree  7 = Strongly agree |
| **Clinic Experience** | | |
| Please rate the length of your visit. | 5-point Likert scale | 1 = Too short  3 = Appropriate length  5 = Too long |
| How easy do you think it will be to return to the clinic and obtain what was prescribed (e.g., follow-up appointments)? | 5-point Likert scale | 1 = Extremely easy  5 = Extremely difficult |
| How easy was it to obtain other Beacon services (e.g., meals, laundry, shower) during your clinic visit? | 5-point Likert scale | 1 = Extremely easy  5 = Extremely difficult |
| **Technology Access** | | |
| On a scale of 1–10, how easy is it for you to access websites? | 10-point Likert scale | 1 = Very difficult  10 = Very easy |
